# The urothelial transcriptomic response to interferon gamma predicts T1 recurrence-free and basal/squamous muscle-invasive bladder cancer survival and better targeted strategies for immune checkpoint blocking

**DOI:** 10.1101/2022.02.04.22270000

**Authors:** Simon C. Baker, Andrew S. Mason, Raphael G. Slip, Pontus Eriksson, Gottfrid Sjödahl, Ludwik K Trejdosiewicz, Jennifer Southgate

## Abstract

Intravesical Bacillus Calmette-Guérin vaccine (BCG) is an established immunotherapeutic in bladder cancer (BlCa), provoking inflammation leading to tumour-specific immunity. Immune checkpoint blockers such as anti-PD-L1 have potential for enhancing tumour-specific lymphocyte-mediated cytotoxicity in BCG-refractive or advanced disease. In both cases, Interferon-gamma (IFNγ) plays a central role. We investigated the transcriptomic response of normal human urothelium to IFNγ to disentangle mechanisms of BCG and anti-PD-L1 therapy failure.

Exposure of differentiated human urothelium to IFNγ resulted in upregulated MHC class I and class II and *de novo* expression of CXCL9-11 chemokine genes. Normal urothelium expressed only immuno-inhibitory B7 family members: PD-L1 expression was induced by IFNγ, whereas VISTA was expressed constitutively.

A urothelial IFNγ response gene set was derived and used for unsupervised clustering of tumours, which predicted longer recurrence-free survival in non-muscle invasive bladder cancer (NMIBC). In muscle invasive bladder cancer (MIBC), the IFNγ-signature split the basal/squamous consensus subtype, with significantly worse overall survival when weak/absent.

Normal urothelium has few resident lymphocytes. Tumour cell killing requires recruitment and activation of IFNγ-secreting pro-inflammatory/cytotoxic lymphocytes while surmounting both innate (VISTA) and upregulated (PD-L1) inhibitory mechanisms. This study offers supportive evidence for strategies to enhance immunotherapy *via* the IFNγ and VISTA/PD-L1 nexus.

**Patient Summary:** Immunotherapy brings promise of harnessing a patient’s own immune system to seek and destroy malignant cells, but it has yet to deliver widespread clinical benefit. We exposed human urothelium to interferon gamma, a key messenger of the immune system and identified a novel signature of 33 genes that predicted cancers with better outcomes. Our study revealed alternative strategies for targeting checkpoint proteins to improve immunotherapy in the future.

## Manuscript

BCG is widely used in NMIBC therapy as a potent inducer of inflammation, provoking infiltration by IFNγ-secreting proinflammatory leukocytes which become exposed to tumour neoantigens, resulting in tumour-specific immunity [1]. In an orthotopic MB49 mouse BlCa model, IFNγ was critical for intrinsic tumour surveillance and an *Ifng*-knockout rendered BCG ineffective [2].

We report the results of exposing mitotically-quiescent (G0-arrested), *in vitro*-differentiated normal human urothelium from six independent donors to 200 U/mL IFNγ to generate a tissue-specific urothelial response transcriptome. mRNA-sequencing identified 107 genes significantly (*q*<0.05) >2-fold increased by seven-day exposure to IFNγ (Fig. 1A and Supplementary Table 1).

**Fig. 1.**
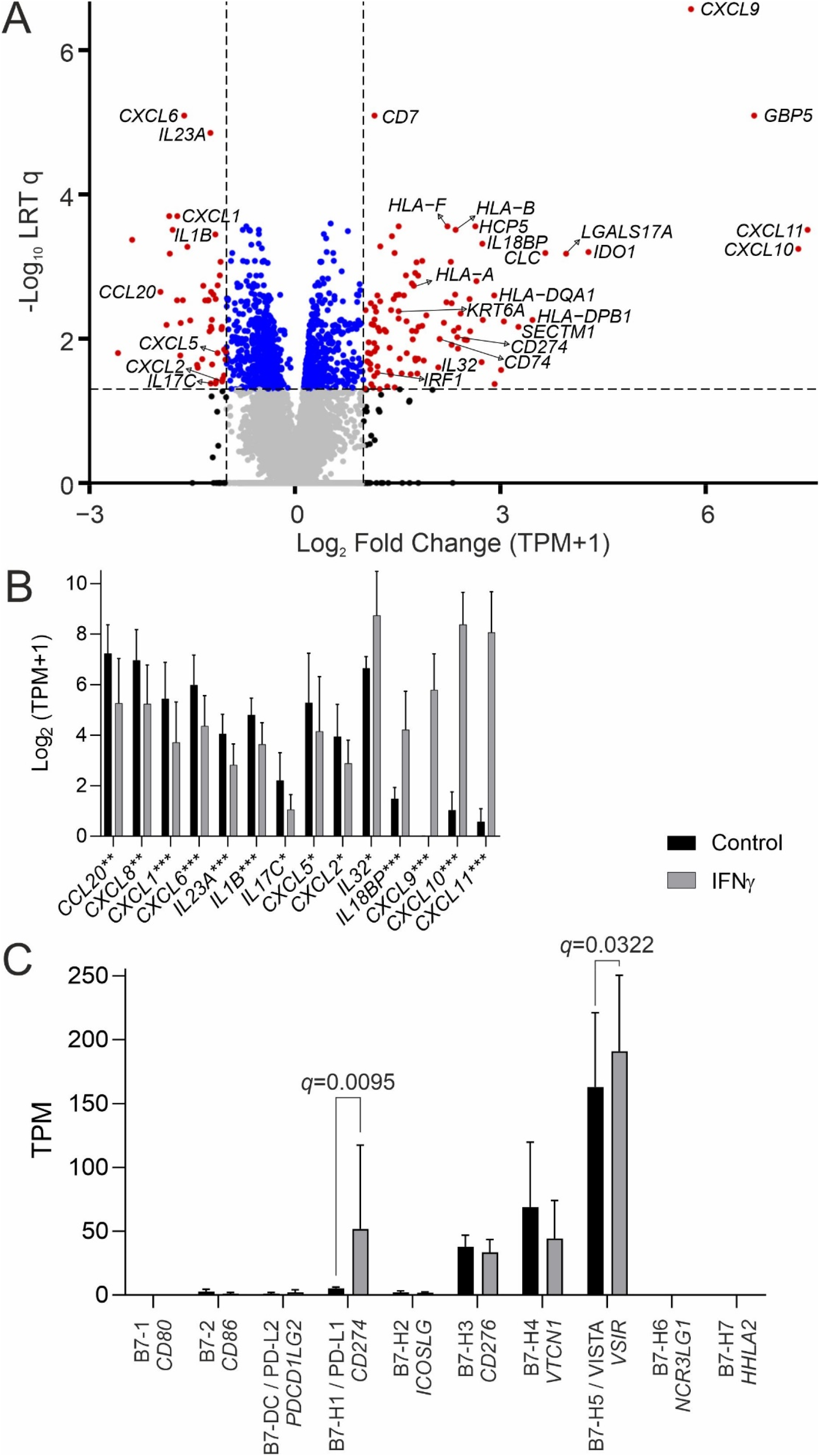
(A) mRNA-sequencing volcano plot for the IFNγ-induced normal urothelial transcriptome. Data is expressed as transcripts per million (TPM) values and significance was assessed using a likelihood ratio test (LRT) to generate Benjamini-Hochberg corrected q-values. The volcano highlights the potential for urothelial-immune signalling via (B) observed shift in chemokine-related gene expression. Stars following gene names indicate >2-fold changes with significance *=q<0.05, **=q<0.01 and ***=q<0.001. (C) Urothelial expression of the family of B7 peripheral membrane protein genes. The MHC co-stimulatory molecules CD80 and CD86 were not expressed. CD274 (“PD-L1”) was significantly induced by IFNγ treatment (mean fold change = 5.2; q=0.0095). VSIR (“VISTA”) was abundantly expressed and significantly induced by IFNγ treatment (mean fold change = 1.2; q=0.0322). For the x-axis labels, the B7-family name and other common names are provided in normal font above the italicised HGNC gene name. Data in all panels is derived from normal human urothelium established from six independent donors.

The IFNγ receptor genes (*IFNGR1* and *IFNGR2*) were abundantly expressed by urothelium (Supplementary Fig. 1). In all six donors, irrespective of haplotype, IFNγ induced gain of human leukocyte antigen (HLA) gene expression associated with both major histocompatibility complex (MHC) class I and class II (Supplementary Fig. 2). Gains in expression of β2-microglobulin (*B2M*), MHC class II invariant chain (CD74) and trans-activator (*CIITA*), as well as the antigen peptide transporters *TAP1* and *TAP2*, suggest an induced role in modulating antigen-presentation to CD4 T cells (Supplementary Fig. 2). However, urothelial cells are unlikely to act as professional antigen presenting cells, because the co-stimulatory B7 family CD80 and CD86 genes were absent and were not inducible by IFNγ(Fig. 1C).

Blocking antibodies against the PD-L1 (CD274) immune checkpoint have been trialled clinically to disinhibit tumour-specific cytotoxic lymphocytes. *CD274* (PD-L1) was strongly induced by IFNγ (Fig. 1C). By contrast, the inhibitory B7 family member *VSIR* (VISTA) was expressed constitutively by normal human urothelium but has to-date been overlooked in the bladder [3]. This indicates that normal urothelium, and likely its malignant counterpart, is a locally immunosuppressive environment, with induction of PD-L1 by IFNγ acting as a negative feedback loop to prevent runaway inflammation.

IFNγ-treatment induced a shift in expression of chemokines and cytokines implicated in immune recruitment, with loss (*CCL20, CXCL8, CXCL1, CXCL6, IL23A, IL1B, IL17C, CXCL5* and *CXCL2*) and gain (*CXCL11, CXCL10, CXCL9, IL18BP, IL32*) of different signalling factors (Fig. 1B). Of particular note, the CXCL9/10/11-CXCR3 axis is a critical regulator of leukocyte migration, differentiation and activation [4], including recruitment of effector T cells. We have shown elsewhere that *CXCL10/11* are highly induced following exposure of normal urothelium to BK virus [5] and were the only genes to be further upregulated, rather than repressed, by treatment with exogenous IFNγ. We therefore suggest that attenuated BK virus could be used to generate a pro-inflammatory, IFNγ-rich local environment akin to that induced by BCG.

Cancer subtyping is usually performed by unsupervised clustering of most statistically informative genes detected as transcripts in extracted tumour tissue; the latter representing a heterogeneous mix of tumour, stromal and immune cells. Whilst able to categorise tumours into subsets by similarity, it is a blunt tool for informing therapy. We hypothesised that using signature gene sets representing tissue-specific pathway responses would reduce “noise” and provide greater therapeutic insight.

The *IFNG* transcript can be detected in cancer cohorts but gives weak sensitivity and, as a diffusible factor, IFNγ can elicit a response in the tumour even when originating from peritumoural leukocytes. We hypothesised that a urothelial IFNγ-response signature would provide a more informative means of detecting IFNγ-signalling in a tumour. We therefore applied a curated urothelial *in vitro* IFNγ-response gene set (Supplementary Table 2) to the unsupervised clustering of BlCa cohorts, yielding an “IFNγ-signature” score.

The IFNγ-signature was significantly correlated with *IFNG* transcript in all four BlCa cohorts analysed (see figure legends for details). In NMIBC from the UROMOL2021 cohort [6], the IFNγ-signature was weak in Class 2a tumours (Supplementary Fig. 3). In T1 tumours (combined from UROMOL2021 [6] and Northwestern Memorial Hospital (NMH) [7] cohorts; Fig. 2A) dichotomised into IFNγ-signature high and low groups, recurrence was 50% more likely in patients whose tumours lacked the IFNγ-signature (Fig. 2B). A Cox proportional hazards regression analysis of the pseudo-continuous unit-length scaled IFNγ-signature scores in T1 tumours predicted a recurrence hazard ratio of 3.174 *p*=0.0230 (Supplementary Tables 3-4).

**Figure 2.**
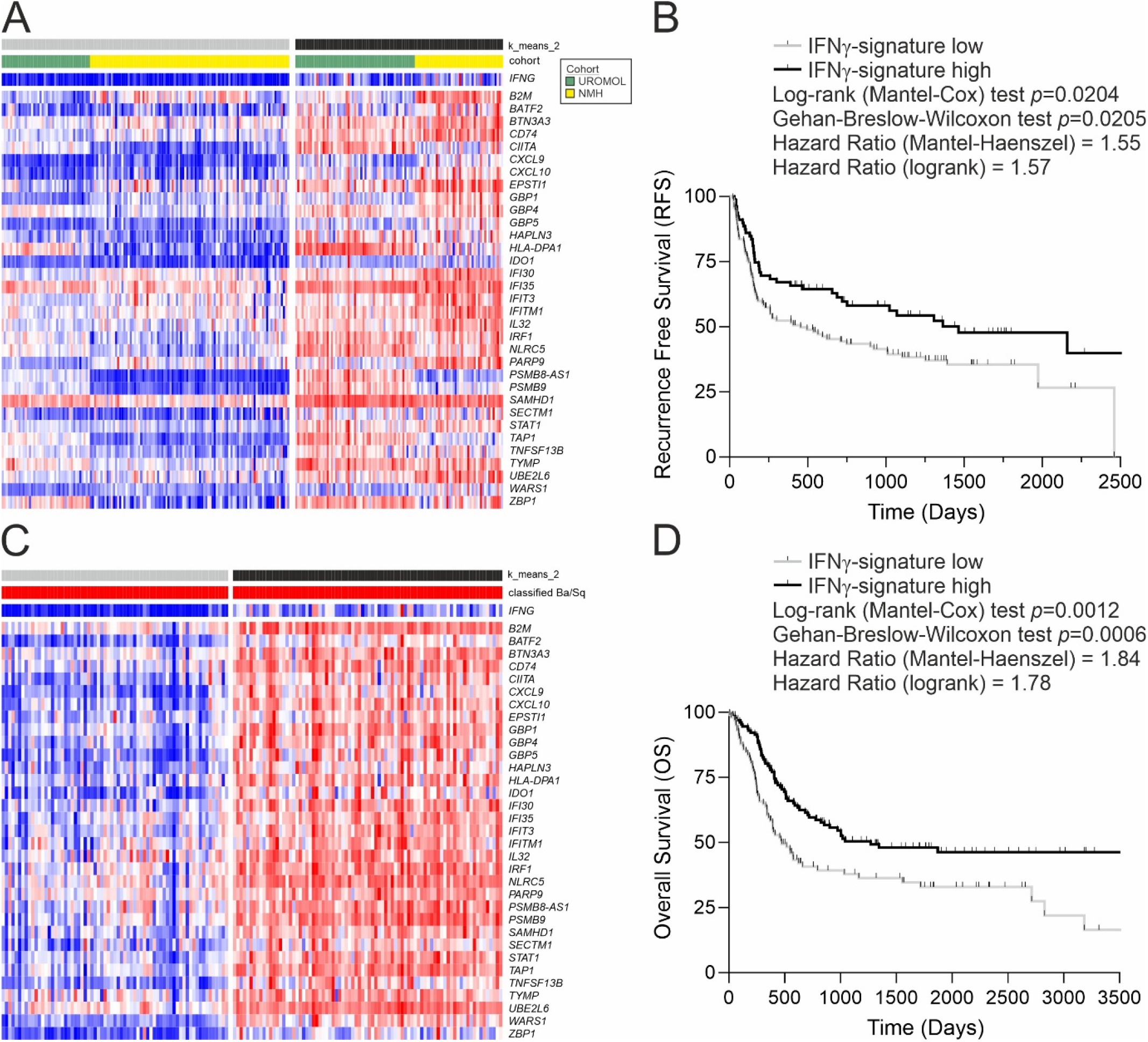
(A) Heatmap and k means clustering based on expression of the IFNγ-signature genes in the T1 tumours of UROMOL2021 and Northwestern Memorial Hospital (NMH) cohorts (n=236). The IFNγ-signature shows a Spearman rank correlation of 0.66 (p=4×10^−31^) with the IFNG gene in T1 tumours. The full UROMOL2021 cohort is shown as Supplemental Fig. 3; [6]). The separate NMH T1 cohort (n=99; [7]) is shown as Supplemental Fig. 4. (B) Kaplan-Meier analysis of IFNγ-signature high and low T1 tumours with survival data from UROMOL2021 and NMH cohorts combined (n=201). The Kaplan-Meier curve is truncated but no events were recorded after the truncation. A Cox proportional hazards regression was also performed using the unit-length scaled IFNγ-signature values from T1 tumours which predicted a recurrence hazard ratio of 3.174 p=0.0230 (Supplementary Table 4). (C) Heatmap for expression of the IFNγ-signature in the Basal/Squamous group of TCGA MIBC tumours (n=151; full cohort is shown as Supplemental Fig. 5; [8]; shown classified in red according to the consensus report [9]). The IFNγ-signature shows a Spearman rank correlation of 0.87 (p=3.85×10^−47^) with the IFNG gene in Ba/Sq TCGA-BLCA tumours. Similar analysis for the Lund MIBC cohort (n=88; [10]) is shown as Supplemental Fig. 6. (D) Kaplan-Meier analysis of the IFNγ-signature in Basal/Squamous tumours from TCGA and Lund MIBC cohorts combined (n=232). The Kaplan-Meier curve is truncated but no events were recorded after the truncation. A Cox proportional hazards regression was also performed using the unit-length scaled IFNγ-signature values from both cohorts which predicted a hazard ratio of 4.846 p=0.0006 (Supplementary Table 7).

Analysis of TCGA-BLCA data [8] revealed that MIBC lacking the IFNγ-signature were most likely to be graded histologically as lymphocyte-negative and of luminal papillary sub-type according to the consensus classification [9] (Supplementary Fig. 5). By contrast, MIBC classed as basal/squamous split into two clear subgroups (Fig. 2C). To examine any difference in outcomes associated with the IFNγ-signature in basal/squamous tumours, the Lund MIBC cohort [10] was analysed in parallel (Supplementary Fig. 6). When dichotomised into IFNγ-signature high and low groups, overall survival for basal/squamous tumours (pooled from TCGA-BLCA and Lund cohorts) was significantly lower in tumours with a weak IFNγ-signature (Fig. 2D). A high survival hazard ratio of 4.846 *p*=0.0006 was predicted by Cox proportional hazards regression based on the pseudo-continuous IFNγ-signature scores from both cohorts (Supplementary Tables 5-7).

Bladder cancer is a disease characterised by high mutational loads derived by different processes, whose historic contributions can be assessed using mutational signatures. IFNγ-signature^high^ basal/squamous MIBC tumours had significantly enriched genomic damage from APOBEC enzymes (Supplementary Fig. 7). Widespread APOBEC damage leads to tumour suppressor loss and creates neoantigens which were significantly more abundant in IFNγ-signature^high^ basal/squamous tumours [8]. In an inflammatory context, neoantigens would induce antigen-specific T cell immunity. Conversely, our data suggest that normal and malignant urothelium would suppress T cell activation through PD-L1/VISTA checkpoint inhibition, thereby inhibiting type-1 anti-tumour immune responses. It seems most probable that a balance becomes established between activated tumoricidal effector cells and regulatory/immunosuppressive elements.

To conclude, this study suggests a reconsideration of the role for IFNγ in motivating immune clearance in non-BCG-responsive NMIBC. The IFNγ-signature^high^ basal/squamous MIBC tumours identified here appear to represent a target for immune checkpoint blockade. However, in the absence of IFNγ, expression of PD-L1 is likely to be too low to provide an effective target. We suggest that BCG therapy would be most effective when combined with blocking of both the innate (VISTA [3]) and inducible (PD-L1) immune checkpoints. We anticipate the urothelium-derived transcriptomic IFNγ-signature will prove a valuable tool for the design of more efficacious immunotherapy protocols.

## Data Availability

All data produced are available online at GSE174244.

https://www.ncbi.nlm.nih.gov/geo/query/acc.cgi

## CRediT Author Statement

Conceptualization – SCB, JS

Methodology – SCB

Software – ASM, SCB, PE, GS

Formal analysis – SCB, ASM

Investigation – SCB, RGS

Data Curation – ASM, SCB, PE, GS

Writing – SCB, LKT, JS

Writing - Review & Editing – SCB, ASM, RGS, PE, GS, LKT, JS

Visualization - SCB

Supervision – SCB, JS

Project administration - SCB

Funding acquisition - JS

## Acknowledgements

This study was funded by York Against Cancer.

