## Supplementary Methods and Figures for "The urothelial transcriptomic response to interferon gamma predicts T1 recurrence-free and basal/squamous muscle-invasive bladder cancer survival and better targeted strategies for immune checkpoint blocking"

### Supplementary Information

**Supplementary Methods**

*Cell culture methods*

Six independent Normal Human Urothelial (NHU) cell lines of finite (non-immortalised) lifespan were used in this study. The cell lines were established as described [1] using anonymous discarded tissue from renal transplant surgery, with UK NHS approval (REC reference 99/095) from the Leeds East Research Ethics Committee. NHU cells were propagated in Keratinocyte Serum-Free Medium containing bovine pituitary extract, recombinant human EGF (KSFM) and additionally supplemented with 30ng/ml cholera toxin. Following expansion, NHU cells were differentiated in the same medium supplemented with 5% adult bovine serum and $CaCl_2$ was added to elevate the [$Ca^{2+}$] from 0.09 to 2 mM, according to published methods [2]. Differentiated cultures of the six independent cell lines were exposed to IFNγ (200U/mL, BioTechne #285-IF) for 7 days and medium was changed every 48 hours. Urothelial mitotic-quiescence and differentiation was largely unaffected by IFNγ-treatment and although there was a significant gain of *KRT6A* expression, there was no significant loss of transitional epithelial markers and no other indicators of squamous change (Supplementary Table 1).

*mRNA Analysis*

Total RNA was collected in TRIzol reagent (Invitrogen) and mRNA-sequencing was performed using an Illumina NovaSeq 6000 generating 150bp paired-end reads (Novogene UK, Cambridge, UK). All mRNA-sequencing data was deposited at GSE174244. Following standard quality control, gene-level expression values in transcripts per million (TPM) were derived against the Gencode v35 human transcriptome using kallisto v0.46.1 [3]. Differentially-expressed genes were identified using the sleuth v0.30.0 [4] implementation of the likelihood ratio test (LRT), accounting for matched genetic backgrounds, generating Benjamini-Hochberg corrected q-values. For volcano plots (performed in R v4.0.4 EnhancedVolcano v1.8.0), fold-change values used a $\log_2$(TPM+1) transformation to reduce the influence of low abundance transcripts. This analysis identified 107 genes that were significantly ($q$<0.05) >2-fold increased by IFNγ and 48 genes whose expression was significantly ($q$<0.05) more than halved by IFNγ-treatment of urothelial tissues (Supplementary Table 1).

*Publicly-Available Bladder Cancer Cohort Data*

Patient data for four publicly-available bladder cancer cohorts was downloaded following instructions in their relevant publications [5-8]. Two cohorts focussed on non-muscle invasive bladder cancer (NMIBC) and from these we extracted only the T1 tumours for further analysis [5, 7]. Two cohorts we of muscle invasive bladder cancer (MIBC) and from these we extracted the tumours transcriptomically classified as the "Basal/Squamous" subtype for further analysis based on the high variability in the IFNγ-Signature that we observed in this group [6, 8, 9]. mRNA-sequencing data was remapped to Gencode v35 human transcriptome using kallisto v0.46.1 where possible [3]. MIBC gene array data from the Lund cohort was utilised as deposited at GSE83586 to generate Combat centered probe fluorescence values [8].

*IFNγ-signature generation*

All $q<0.05$ significantly >2-fold IFNγ-increased genes were extracted from the TCGA-BLCA [6] and UROMOL2021 [7] cohorts of MIBC and NMIBC, respectively. Genes regulated by IFNγ *in vitro* can be regulated predominantly by other mechanisms in tissues and to refine the signature, Spearman correlation matrices were calculated to compare all genes with one another. The median Spearman Rho for each gene in comparison with all others was calculated in each cohort. The median Spearman Rho values for the TCGA-BLCA and UROMOL2021 cohorts were averaged and only genes with a value greater than 0.5 were retained (n=33; gene list in Supplementary Table 2) to generate a transcriptomic classifier relevant to both NMIBC and MIBC, where all genes were >2-fold increased by IFNγ *in vitro* and closely related to one another in tumours.

Initial evaluation of the IFNγ-signature in tumours was performed on $\log_2$(TPM+1) data using hierarchical clustering (based on Euclidean distance and complete linkage. This approach highlights the relationship between samples in the full cohorts of NMIBC UROMOL2021 [7] (Supplementary Fig. 3) and MIBC TCGA-BLCA [6] (Supplementary Fig. 5).

*IFNγ-signature analysis*

mRNAseq data expressed as TPMs for tumours from the NMIBC cohorts was combined to create one large cohort for analysis. For the MIBC cohorts, to allow integration of data acquired by different methods (TCGA-BLCA mRNA-sequencing [6] with the Lund gene array [8] data) each cohort was clustered separately into IFNγ-signature high and low groups. Classification of tumours was performed on $\log_2$(TPM+1) data using Euclidean distance k means clustering into two groups of T1 NMIBC tumours (from the UROMOL2021 [7] and Northwestern Memorial Hospital [5] cohorts) and Basal/Squamous classified [9] MIBC tumours (TCGA-BLCA [6]). Classification of Basal/Squamous MIBC tumours form the Lund [8] cohort was performed using Euclidean distance k means clustering into two groups on log-transformed Combat centered probe fluorescence.

A single-value IFNγ-signature score was derived by unit-length scaling the $log_2$(TPM+1) data (or log-transformed Combat centered probe fluorescence for the Lund cohort [8]) for each gene. Unit-length scaled data for the genes in the signature were then summed on a per patient basis before being re-scaled (again 0-1). This derived a single value IFNγ-signature score that could be ranked to generate Spearman Rho values in comparison with ranked gene expression values (eg *IFNG*) in the various cohorts. The single-value IFNγ-signature scores for the combined T1, TCGA-BLCA and Lund cohorts are provided in Supplementary Tables 3, 5 and 6, respectively.

Survival analysis was performed in Prism v9.3.1 (Graphpad). T1 NMIBC tumours and Basal/Squamous classified MIBC tumours from the different cohorts were pooled to support Kaplan-Meier NMIBC Recurrence Free Survival (RFS) and MIBC Overall Survival (OS) analysis, respectively. Statistical significance of the difference between Kaplan-Meier curves for IFNγ-signature high and low groups was performed using Mantel-Cox and Gehan-Breslow-Wilcoxon tests, with hazard ratios calculated using both Mantel-Haenszel and log rank approaches. A Cox proportional hazards regression analysis was performed using the pseudo-continuous unit-length scaled IFNγ-signature scores to demonstrate that the effects of IFNγ-signalling persist without dichotomising the tumours.

*TCGA-BLCA mutational signature analysis*

TCGA-BLCA legacy mutation data [6] was downloaded from cBioPortal [10]. The single base substitution (SBS) signature for all tumours was analysed using the "Catalog" input for the "signal" workflow for mutational signature analysis by comparison against the Catalogue of Somatic Mutations In Cancer "COSMIC" reference SBS signatures common in bladder tumours (namely SBS2, SBS4, SBS5 and SBS13; https://signal.mutationalsignatures.com/ [11]).

**Supplementary Figures**

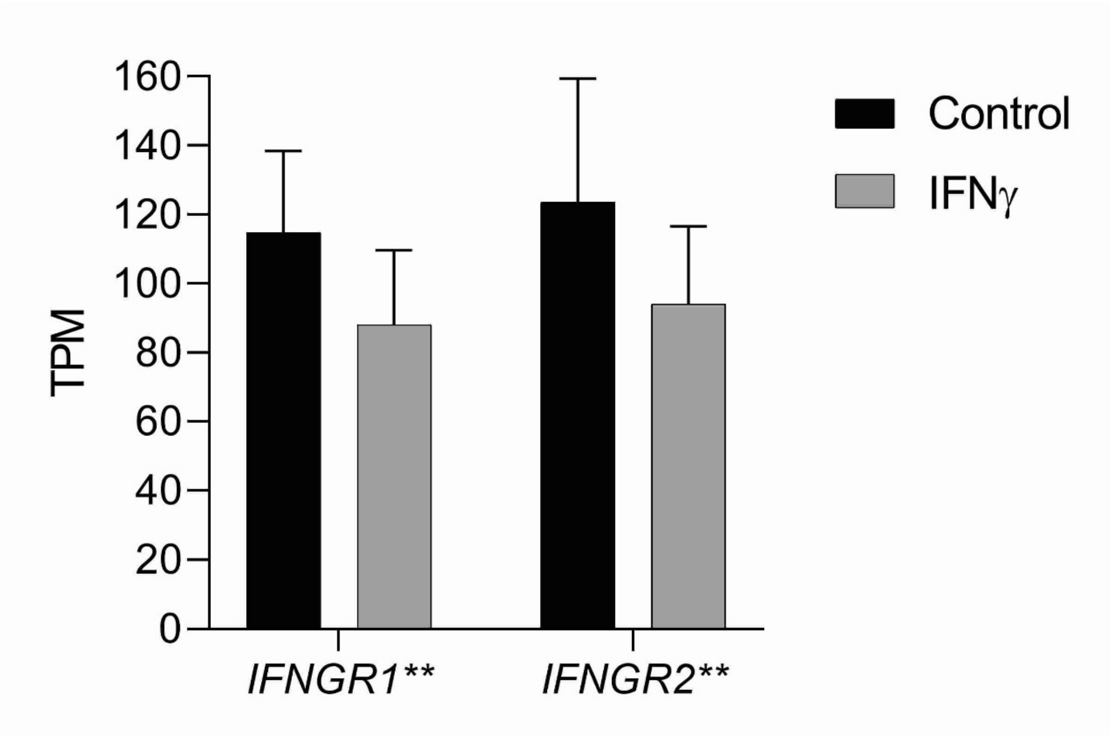


*Supplementary Figure 1 – Urothelial expression of both IFN$\gamma$ receptor isoform genes was high but significantly reduced by IFN$\gamma$ treatment. Mean $log_2$ fold changes were -0.39 and -0.38 for IFNGR1 and IFNGR2, respectively. Stars following gene names indicate significance **=p<0.01.*

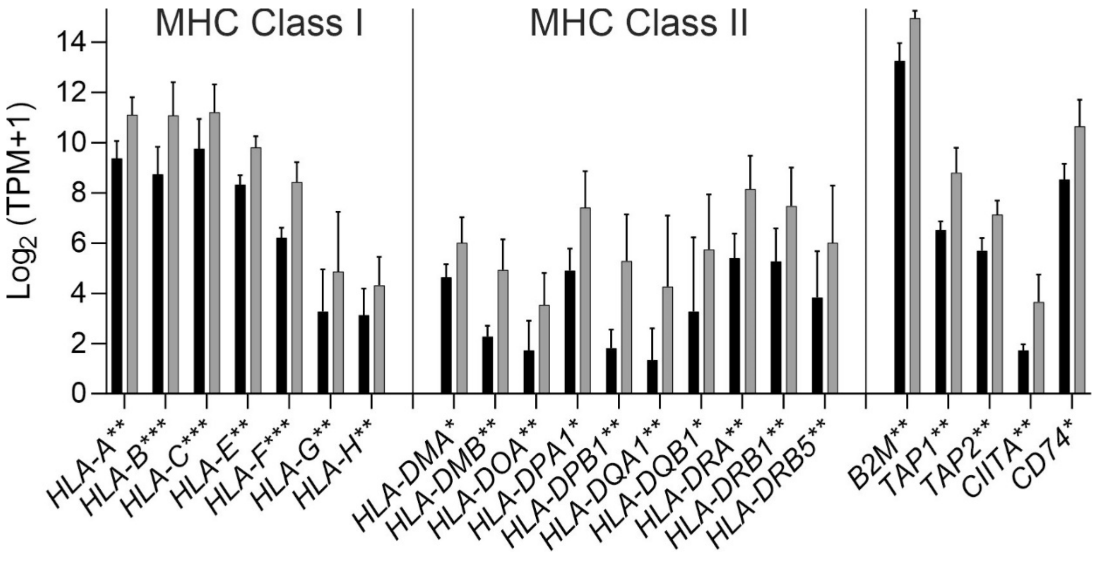


*Supplementary Figure 2 – Gain in expression of a broad range of human leukocyte antigen (HLA) genes. This study used cell derived from six independent donors. Stars following gene names indicate >2-fold changes with significance *=q<0.05, **=q<0.01 and ***=q<0.001.*

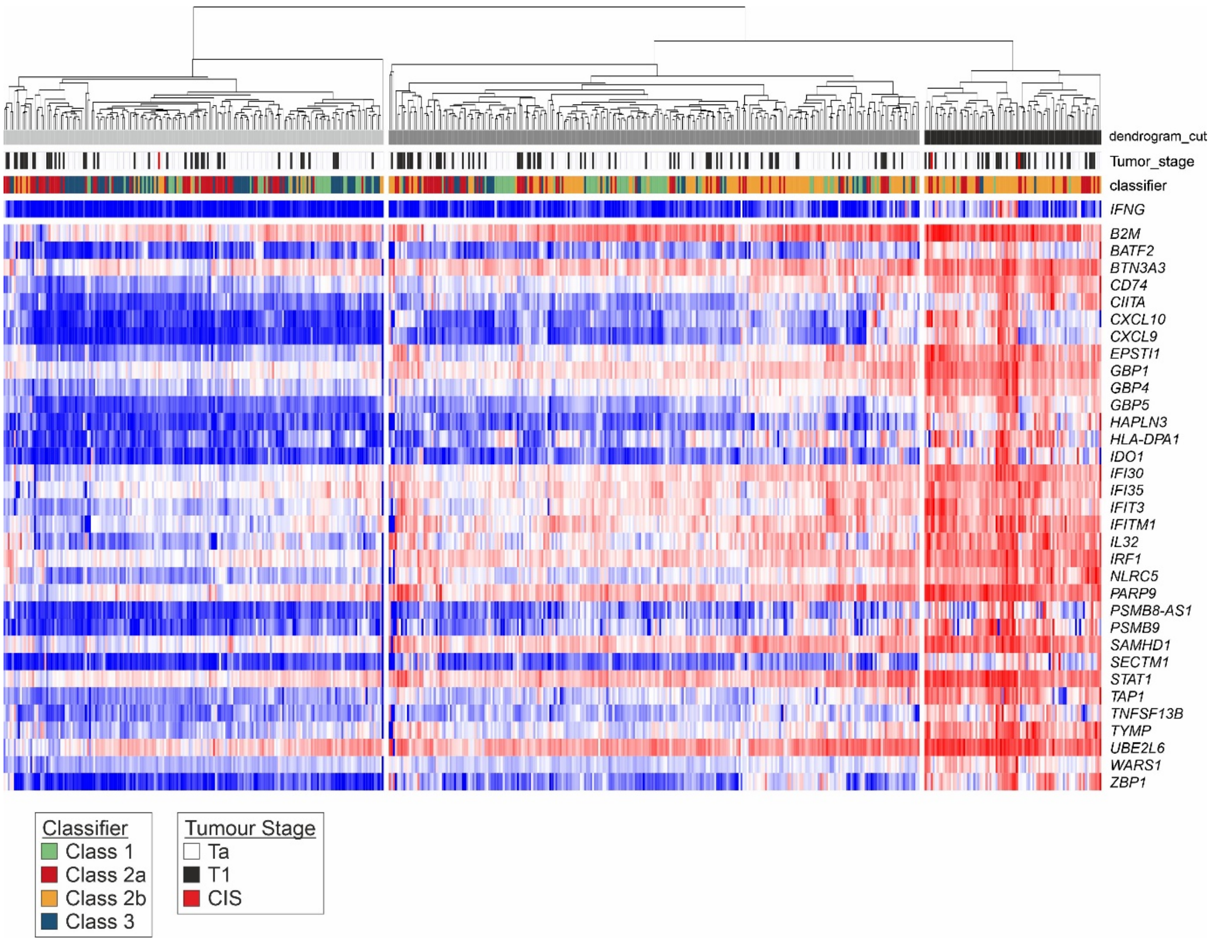


*Supplementary Figure 3 – The full UROMOL2021 NMIBC cohort (n=535; [7]) expression of the IFNγ-signature genes. Tumours were split using hierarchical clustering based on Euclidean distance with complete linkage. The IFNG gene is not part of the signature but is included above to show the lack of sensitivity when relying on IFNG transcript abundance alone. IFNG transcript abundance was significantly correlated with the IFNγ-signature (Spearman Rho=0.57; p=1.02x10$^{-46}$). Tumours are coloured according to the 2021 classification into four subtypes (Class 1, Class 2a, Class 2b and Class 3) [7]. The heatmap shows enrichment of IFNγ responsive gene expression in a subset of the Class 2b group of NMIBC. Lindskrog et al. identified Class 2b as being the most immune-infiltrated tumours [7]. Class 2a tumours were predominantly IFNγ-signature$^{low}$ and had the highest recurrence rate in the original publication [7].*

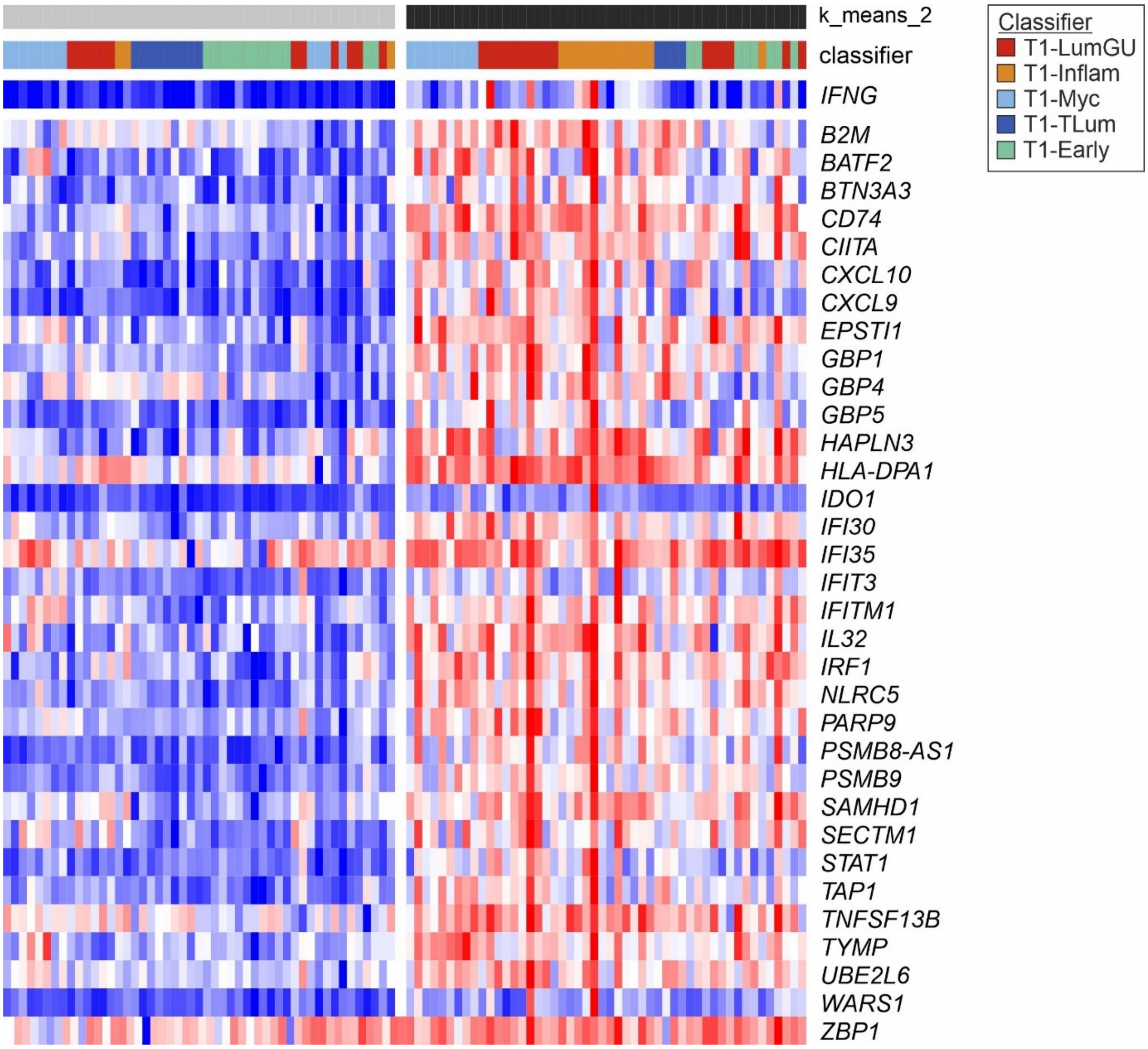


*Supplementary Figure 4 – Heatmap and k means clustering based on expression of the IFN$\gamma$-signature in the T1 tumours of the Northwestern Memorial Hospital (NMH) cohort (n=99; shown with the classifier from the original report [5]). The IFN$\gamma$-signature shows a Spearman rank correlation of 0.67 (p=$1.66\text{x}10^{-14}$) with the IFNG gene in this cohort.*

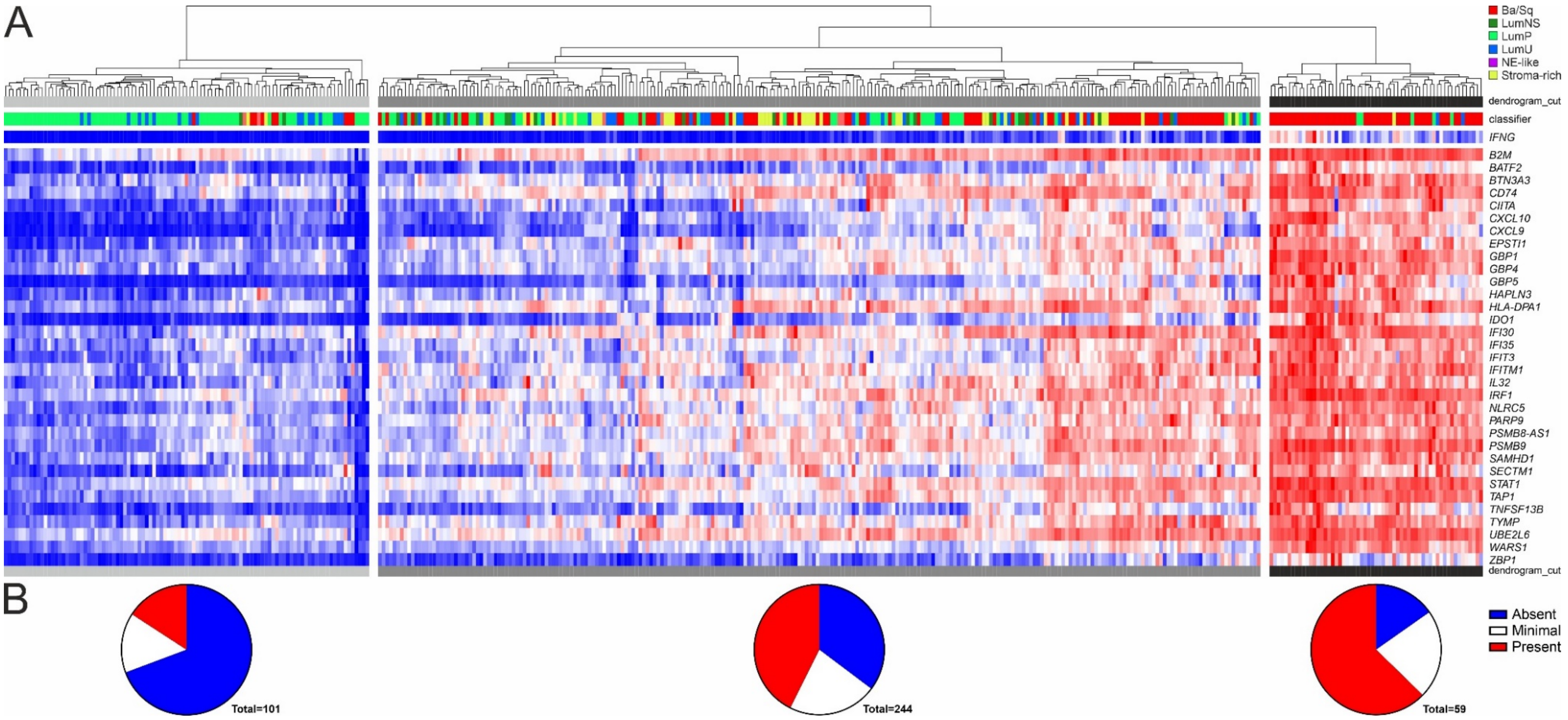


*Supplementary Figure 5 – (A) The full TCGA-BLCA MIBC cohort (n=404; [6]) expression of the IFNγ-signature genes. Tumours were split using hierarchical clustering based on Euclidean distance with complete linkage. The IFNG gene itself is not part of the signature and has a limited sensitivity for detecting IFNγ signalling; however, it does show significant correlation with the IFNγ-signature (Spearman Rho=0.83; $p=2.86x10^{-102}$). Tumours are coloured according to the 2019 consensus classification into six subtypes (Basal/Squamous, Luminal Non-specified, Luminal Papillary, Luminal Unstable, Neuroendocrine-like and Stroma-rich) [9]. The heatmap shows diversity in the IFNγ-signature within the Basal/Squamous group of MIBC and so these tumours were evaluated further (Figure 2C). (B) Histological grading of lymphocyte invasion of TCGA-BLCA tumours, showing significant (Chi square=56.63; df=4; $p=1.48x10^{-11}$) differences, including that IFNγ-signature low tumours were more likely to contain no lymphocytes.*

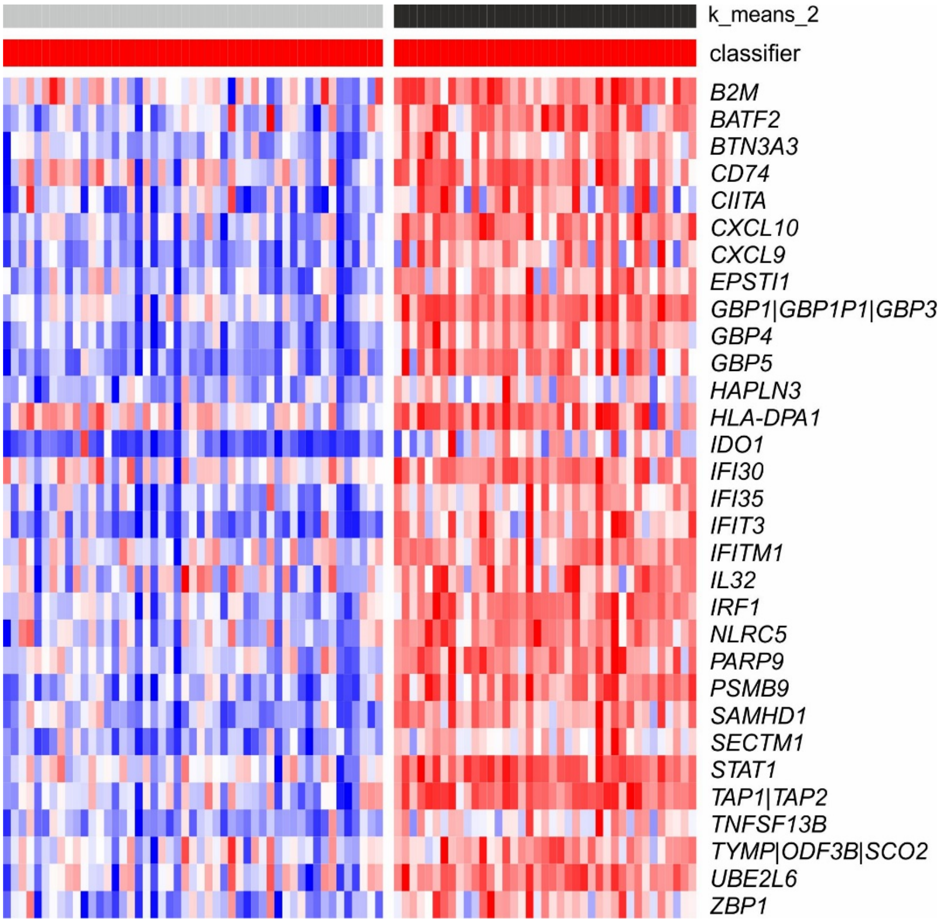


*Supplementary Figure 6 – Basal/Squamous classified tumours from the Lund MIBC cohort (n=88) showing expression of the IFNγ-signature genes (where present on the gene arrays employed). Multiple gene names indicate the gene array probes bind multiple targets. There was no specific probe for the IFNG gene and so correlation with the signature could not be calculated in this cohort.*

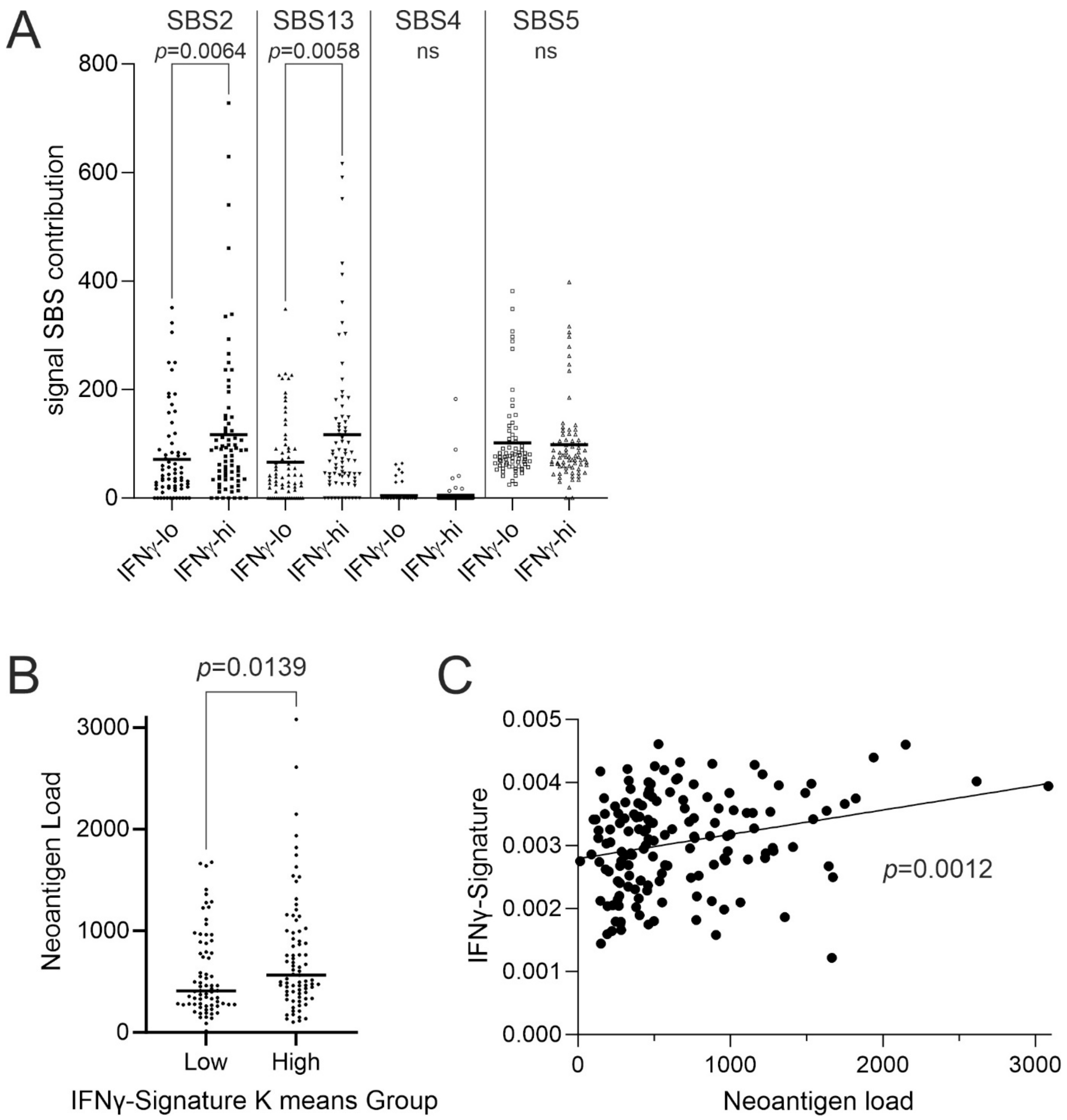


*Supplementary Figure 7 – Basal/Squamous tumours of TCGA-BLCA have paired exome-sequencing which allows the analysis of mutations. No significant over-mutation of individual genes was observed in association with the IFNγ-signature. (A) However, APOBEC-driven mutational processes had been significantly more active (as denoted by significant enrichment of single base substitution signatures "SBS2" and "SBS13") in tumours that ultimately had higher IFNγ-signature scores. It was not the case that all mutational processes were more active in forming tumours with a higher IFNγ-signature, as SBS4 and SBS5 were*

*not enriched. Significance was analysed by Mann Whitney U test. ns = no significant difference.*

*In silico analysis of the likely effects of mutations on neoantigen load in tumours was previously described by TCGA consortium* [6]*. (B) Analysis of Basal/Squamous tumours of TCGA-BLCA cohort showed a significantly higher predicted neoantigen load in the context of higher IFN$\gamma$-signalling (p=0.0139). (C) Furthermore, linear regression analysis confirmed a significant relationship between predicted neoantigen load and IFN$\gamma$-signature score (p=0.0012).*
